# Network-Based Stratification Refines Stratification of Intermediate-Risk Acute Myeloid Leukemia Samples

**DOI:** 10.1101/2025.07.04.25330879

**Authors:** Ankita Srivastava, Joseph Saad, Philipp Sergeev, Markus Vähä-Koskela, H. Joachim H. Deeg, Jerald P. Radich, Kernyu Park, Caroline A. Heckman, Janghee Woo

**Author notes:** Corresponding authors: Dr. Janghee Woo, MD, PhD, Winship Cancer Institute, Department of Hematology and Medical Oncology Emory University School of Medicine, 100 Woodruff Circle, Atlanta, GA 30322 USA, Email address; Dr. Caroline A. Heckman, Institute for Molecular Medicine Finland - 14 FIMM, University of Helsinki, P.O. Box20 (Tukholmankatu 8), 00014 University of Helsinki, 15 Finland. Indicate equal contribution.

## Abstract

The European LeukemiaNet (ELN) risk stratification of acute myeloid leukemia (AML) uses genetic and molecular markers to categorize patients. However, disease heterogeneity, particularly in the intermediate-risk group, complicates stratification. Ideker et al. developed the Network-Based Stratification (NBS) method, combining protein network analysis and mutation profiling via machine learning. We applied NBS to intermediate-risk AML patients to refine prognosis and identify distinct molecular subtypes compared to the 2022 ELN scheme. We selected 170 intermediate-risk AML patients based on the 2022 ELN classification from TCGA (n=58), BEAT AML (n=87), and FIMM (n=25) datasets. Using NBS, we analyzed 3,108 genes from WGS or WES data, mapping them onto a cancer-specific protein network for clustering based on network-propagated mutation profiles. We conducted 200 iterations of sub-sampling, considering patients with at least 3 mutated genes and using consensus clustering for robust stratification, assessing associations with clinical and transcriptomic features. NBS identified five distinct molecular subgroups characterized by unique mutation patterns: *IDH1*-dominant (Cluster 1), *DNMT3A*-dominant (Cluster 2), low-frequency multi-mutated (Cluster 3), *FLT3*/*NPM1*/*DNMT3A* co-mutated (Cluster 4), and *FLT3*-dominant (Cluster 5). Cluster 4 showed significantly worse overall survival (HR = 1.81; p = 0.05). In addition, *ex vivo* drug sensitivity and transcriptomic analyses revealed significant variation in therapeutic response and pathway activation across clusters. These findings underscore the power of machine learning–driven approaches like NBS to uncover hidden molecular structure within intermediate-risk AML groups, enabling more precise prognostication and potentially informing personalized therapeutic strategies.

**Key points:**

1. ML-based NBS stratification reveals distinct subgroups within intermediate-risk AML with unique molecular and clinical profiles.
2. AML with *NPM1*/*FLT3*-ITD/*DNMT3A* mutations define a high-risk group with distinct drug sensitivities, including FLT3 inhibitors.

## INTRODUCTION

Acute myeloid leukemia (AML) encompasses a heterogeneous group of diseases, as reflected in the molecular profiles characterized by high-throughput genomic technologies.^1^ This heterogeneity complicates risk stratification based on the subset of identified cytogenetic abnormalities and causal mutations. Therefore, it remains challenging to use cytogenetics to predict AML patient response to induction chemotherapy and survival outcomes, particularly among AML patients classified as normal or intermediate risk.^2^ Also, the recent development of newer targeted therapies has complicated risk stratification to predict clinical outcomes despite the guidelines from the European LeukemiaNet (ELN) ^3–5^.

In an effort to better differentiate these cases, several studies have linked gene expression profiles and epigenetic alterations with treatment outcomes.^6–8^ However, clinical translation of these findings has proven difficult, perhaps because gene expression is affected by multiple factors, including sample processing in clinical settings.^9–11^ For example, standard blood collection procedures rapidly change the transcriptional and post-transcriptional landscapes of hematopoietic cells and introduce potentially confounding effects of technical artifacts in cancer genomics data. ^10^ On the other hand, routine clinical care now often incorporates genomic profiling in myeloid malignancies to identify well-defined common mutations. Unlike transcriptomes and methylomes of a given sample, mutation profiles based on DNA sequencing offer more robust genetic characterization of individual cancers.

Cancer can arise from mutations in single genes, which lead to complicated and interconnected disruptions in molecular pathways. Therefore, similar cancer types may result from mutations that affect different genes yet are involved in a common pathway. In addition, the sequence of mutation acquisition may influence pathologic and clinical behavior of the cancer.^12^ To overcome this limitation in individual mutation analysis^13^, Ideker and colleagues^14,15^ developed a machine learning (ML) network-based method, called Network Based Stratification (NBS), that incorporates functional network analysis as well as somatic mutation profiling. The cancer-specific network including AML-associated genetic alterations was built and validated.^15^ NBS allows clustering of somatic mutation profiles into robust tumor subtypes that have distinct alterations in molecular pathways, and NBS-generated clusters have a strong association with clinical outcomes in various cancers.

By combining functional pathway analysis with mutation profiling, we applied the NBS method alongside a cancer-specific interaction network to a well-curated dataset of patients classified as intermediate-risk AML under current ELN guidelines ^3^. This approach successfully identified a high-risk subset within the intermediate-risk group, characterized by distinct molecular pathway activation and differential drug response profiles. These findings support the utility of ML–based stratification in uncovering clinically significant subgroups that are not identifiable through existing ELN risk criteria, highlighting its potential as both a prognostic tool and a novel biomarker for guiding personalized AML treatment.

## METHODS

### Study cohorts

Patients with intermediate-risk AML as defined by the 2022 ELN genetic risk stratification at the time of diagnosis^3^, were selected from studies by TCGA^1^, BEAT dataset ^16^ and the Institute for Molecular Medicine Finland (FIMM) at the University of Helsinki^17,18^. To maintain cohort homogeneity and avoid confounding effects from prior treatments, only samples collected at the time of diagnosis were included for stratification within the intermediate-risk group among these patients. A total of 87 patients from Tyner et al. were used for the analysis and combined with 58 patients with intermediate-risk AML from the TCGA AML cohort^1^ (Supplementary Figure 1A). Bone marrow mononuclear cells were obtained from the Helsinki University Hospital Comprehensive Cancer Center after informed consent (permit numbers 239/13/03/00/2010, 303/13/03/01/2011, Helsinki University Hospital Ethics Committee) and in compliance with the Declaration of Helsinki. A total of 25 patients from Helsinki University Hospital were analyzed. *Ex vivo* drug sensitivity and resistance testing was performed on the FIMM samples as we have reported previously^17^.

### NBS and transcriptome analysis

NBS was applied following previously reported procedures.^14,15^ In total, 3,108 genes from either whole-genome or whole-exome sequencing were analyzed and applied to the protein interaction pathway map in the previously validated cancer-specific network, spreading the signal to other functionally related genes in the network space. This step enabled clustering of patients based on similarity of network-propagated mutation profiles. We then performed 200 iterations of sub-sampling and consensus clustering for robust and reproducible stratification. Propagated mutation scores, which quantitatively represent the degree of alteration in affected genes and neighborhoods, were used to identify molecular pathways differentially altered in assigned clusters, as previously reported.

NBS-assigned clusters were tested for associations with survival outcomes by Kaplan-Meier analysis (log-rank test) using R version 4.5.0 with survival package v3.8.3^19^. Differentially expressed transcripts between clusters were inferred using DESeq2 (v1.48.0)^20^. An FDR cut-off of 0.05 and |LogFC| ≥ 2 was used to select differentially expressed genes. Pathway enrichment analysis was performed using the fast gene set enrichment analysis (fgsea) method (v1.33.4)^21,22^. Ranked gene statistics were derived from differential expression analysis, and hallmark pathways were used for enrichment. Pathways with an adjusted p-value (padj) < 0.01 were considered significantly enriched. The biological significance of differentially expressed genes was assessed using Gene Set Enrichment Analysis (GSEA) and Gene Ontology (GO) biological processes assessed using clusterProfiler (v4.15.1)^22^. These analyses were performed for each cluster, using other clusters as the reference. Subsequently, we performed a similar analysis between Cluster 4 *vs*. Cluster 5 using the latter as a reference.

## RESULTS

### Network-based stratification identifies genetic subtypes in intermediate-risk AML

Data from 170 patients with intermediate-risk AML, classified according to the 2022 ELN genetic risk stratification at diagnosis^3^, were analyzed using the NBS approach. These samples were sourced from TCGA^1^, BEAT AMLt ^16^, and FIMM datasets^17,18^. We used the human cancer reference network as the previously validated protein-protein functional pathway network, which consisted of 18,487 nodes of protein interaction.^15^ To improve clustering stability, we further restricted our study to patients with at least three mutated genes in the network. The NBS approach identified five distinct AML subgroups: Cluster 1 with 21 patients, Cluster 2 with 34 patients, Cluster 3 with 67 patients, Cluster 4 with 23 patients, and Cluster 5 with 25 patients (Figure 1A and B). Notably, samples from the three sources were evenly distributed across the clusters, except that none of the samples from TCGA were present in Cluster 4 (Figure 1A). Although mutations in *DNMT3A* appeared across all clusters, known leukemia-associated mutations were predominantly enriched in specific clusters. For example, *IDH1* was most frequently mutated in Cluster 1 (100%), *DNMT3A* mutations were highly enriched in Cluster 2 (97.1%), *NPM1* and *FLT3* mutations co-occurred at full frequency in Cluster 4 (100%), and *FLT3* was the dominant mutation in cluster 5 (96%). On the other hand, Cluster 3 did not display a single dominant mutation. Instead, several gene mutations were observed at comparable but low frequencies, including *WT1* (22.4%), *IDH2* (13.4%), *NRAS* (13.4%), *DNMT3A* (11.9%), and *FLT3* (11.9%). This distribution indicated that Cluster 3 was characterized by a heterogeneous mutational profile, marked by various clonal hematopoiesis (CH)-associated mutations but lacking a dominant driver mutation, in contrast to other clusters enriched for mutations such as *IDH1*, *DNMT3A*, *FLT3*, or co-occurring *NPM1* and *FLT3*. Notably, *DNMT3A* was present in 57.1% of Cluster 1, 97.1% of Cluster 2, 11.9% of Cluster 3, 65.2% of Cluster 4, and 20% of Cluster 5, highlighting its widespread but variable involvement across subtypes (Figures 1C and D).

**Figure 1.**
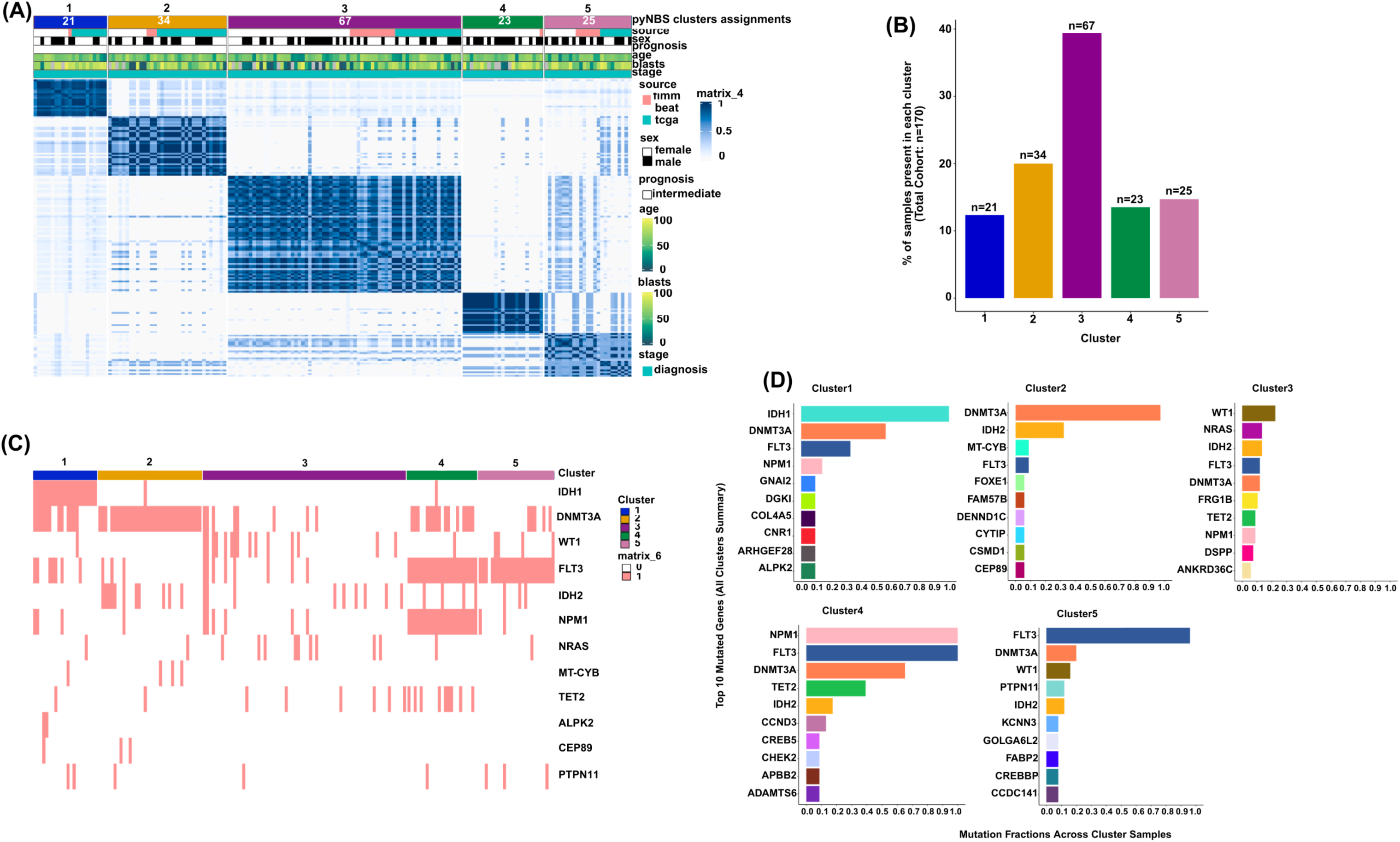
Network-based patient clustering reveals genetic subtypes in intermediate-risk AML. **A)** NBS identifies five distinct clusters in our datasets, with all three datasets (TCGA, FIMM, and BEAT AML). **B)** Number of patients in each cluster assignment **C)** Heatmap of most mutated genes among the 5 clusters. A pink bar indicates a mutant sample, while a white bar indicates a wild-type (WT) sample. For example, Cluster 1 is characterized by *IDH1* mutants with a high fraction for co-occurring *DNMT3A* mutants. Cluster 3 samples consist of many mutually exclusive gene mutations. **D)** Mutation frequencies of the 10 most mutated genes in each cluster. Cluster 4 is characterized by a high rate of co-occurring *NPM1*, *FLT3*, and D*NMT3A* mutations.

### NBS-defined AML subtypes reveal distinct clinical profiles and outcomes

We tested whether pathway-inferred mutation clustering by NBS can identify a subgroup of high-risk patients or varied clinical and therapeutic responses within the intermediate-risk AML. We systematically assessed a comprehensive set of clinical and pathological parameters across all patients in the cohort, including gender, age at diagnosis, bone marrow blast percentage, cytogenetic abnormalities, AML subtype (de novo, secondary, or therapy-related), treatment type (such as allogeneic transplantation), clinical outcomes following induction chemotherapy (7+3 or variants), and overall survival. These parameters were compared across the NBS-defined clusters to identify subgroup-specific patterns. The numbers of mutations in each cluster were not significantly different, but there was a trend of slightly lower mutation burden in Cluster 3 and 4 (Figure 2A). Additionally, there was no significant difference in age at diagnosis or sex distribution among the clusters. Notably, the proportion of male samples (65.2%) in cluster 4 was slightly higher compared to the other clusters (Supplementary Figure 1B and C). We examined *FLT3* mutation distribution across clusters and found that Cluster 4 uniquely harbored *FLT3*-ITD in all samples (Figure 2B). *FLT3*-ITD is well established to counteract the favorable prognosis typically conferred by *NPM1* mutation, resulting in poor outcomes, high white blood cell counts, and increased blast percentage^23–25^. Consistent with this, we observed a trend toward higher bone marrow blast percentage in cluster 4, although not statistically significant (Figure 2C). This finding was supported by a hazard ratio (HR) of 1.81 for Cluster 4 compared to all other clusters (95% confidence interval: 1.0–3.28; p = 0.05), indicating a marginally significant association with poorer outcomes (Figure 2F and 2G). In contrast, other clusters (1, 2, 3, 5) exhibited heterogeneous *FLT3* mutation profiles (ITD, TKD, or both), likely contributing to greater clinical variability within these groups (Figure 2B).

**Figure 2.**
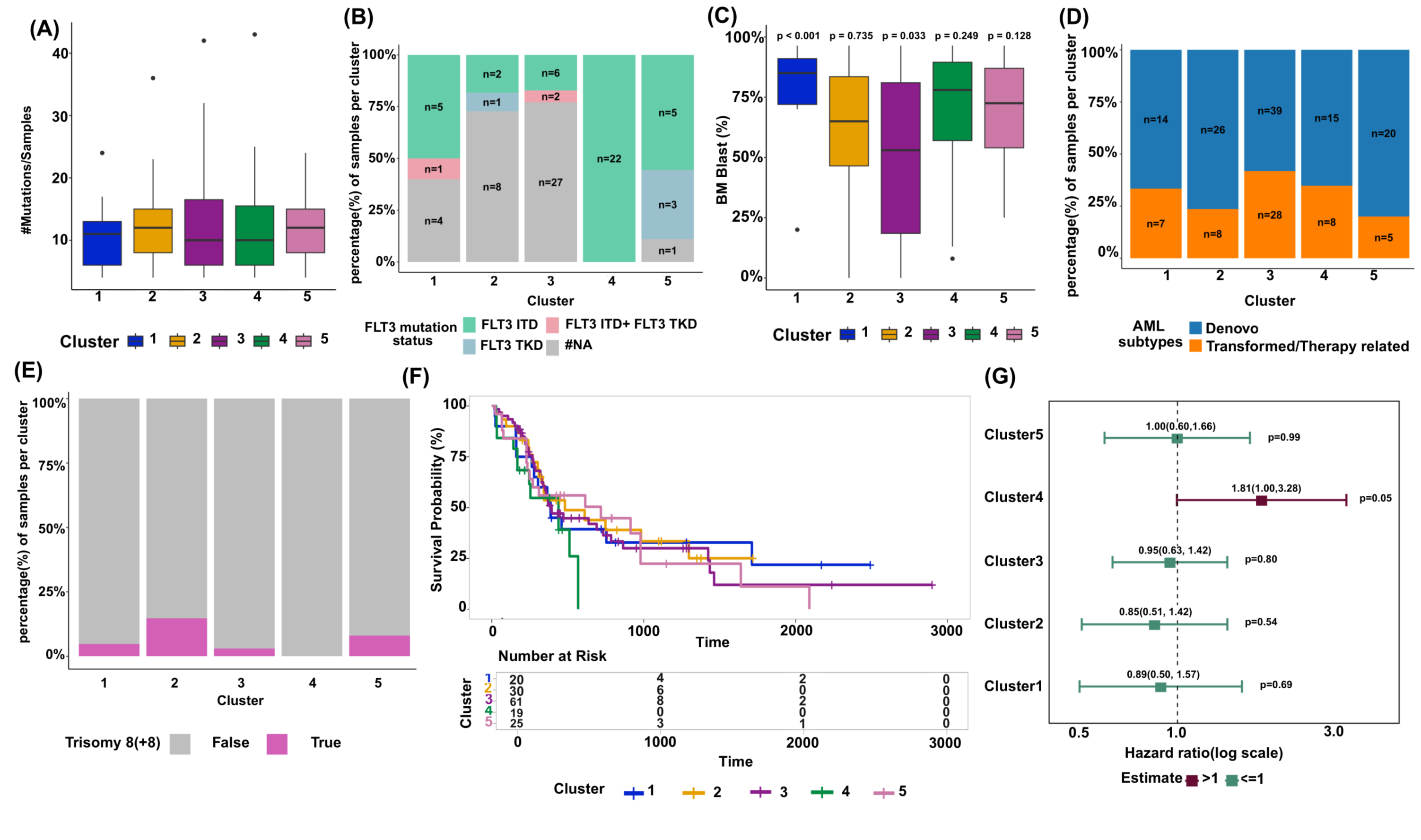
Molecular and clinical associations of NBS-defined AML subtypes reveal distinct clinical profiles and outcomes. **A)** Mutation burden by cluster assignment. **B)** *FLT3* mutation status in BEAT AML samples across five distinct clusters. Patients were grouped in 4 categories according to their mutation status: *FLT3*-TKD, *FLT3*-ITD, *FLT3* Other (non-TKD, non-ITD) and *FLT3* WT. All samples in Cluster 4 exhibited the *FLT3*-ITD mutation. **C)** Comparison of bone marrow blast percentages (BM blast %) across five distinct clusters. T-test p-values at the top of each boxplot show significance of each cluster vs. all others combined. Blast BM blast % of Custer 1 was significantly higher as compared to other clusters. **D)** Proportions of AML subtypes across cluster assignment: de Novo and transformed/therapy related. Cluster 3 has the highest proportions of transformed/therapy-related AML subtype samples. **E)** Percentage of samples that exhibit trisomy 8 in each cluster. Trisomy 8 was detected in 14.7% of samples in Cluster 2, which is highest in proportions among the 5 clusters. **F)** Kaplan-Meier Overall Survival Probability by cluster. **G)** Hazard ratio plot of each cluster. Hazard ratio (HR) (Cluster 4 vs. all other clusters) was 1.81 (95% confidence interval (CI): 1.0–3.28; p = 0.05).

The blast percentage was significantly different between the clusters. Cluster 1, dominated by *IDH1* mutations, showed a significantly higher (p < 0.001) bone marrow blast percentage compared to other groups, which is consistent with reports that *IDH1*-mutated AML can present with elevated blast counts.^26^ In contrast, Cluster 3, which lacks a dominant mutation and has a heterogeneous mutational profile, marked by various CH mutations, exhibited the lowest blast percentage (p=0.033) among the five groups (Figure 2C). This suggests that the absence of a strong driver mutation and the presence of genetic heterogeneity may result in a distinct disease phenotype, with lower blast burden, as seen in myelodysplastic syndrome (MDS). Patients in Clusters 2 (85.7%) and 5 (77.8%) responded to induction chemotherapy better than those in other clusters, whereas 42.9% of patients in cluster 1 were refractory. Clusters 3 and 4 exhibited a moderate treatment response (Supplementary Figure 1D). Patients in Clusters 2 (76.5%) and 5 (80.0%) also had the highest proportions of de novo AML cases, suggesting these clusters are predominantly composed of patients with primary disease. In contrast, Clusters 3 (41.8%) show the highest proportions of transformed or therapy-related AML, indicating a greater presence of secondary or treatment-associated disease (Figure 2D), consistent with their mutational profile and clinical features. Cluster 1 and 4, represent a moderately mixed subtype distribution (Figure 2D). Clusters 2 and 5 may represent biologically favorable subgroups with better therapeutic outcomes, despite showing relatively elevated frequencies of trisomy 8 at 14.7% and 8.0%, respectively, which is often associated with chemoresistance^27^ (Figure 1E). Stem cell transplantation was performed at the lowest frequency in Cluster 2 (23.5%) and at the highest in Cluster 4 (39.1%). However, the lower overall survival observed in Cluster 4 despite the higher transplantation rate suggests that additional high-risk features within this cluster may outweigh the potential benefits of transplantation (Supplementary Figure 1E).

### NBS-Defined AML Subtypes Show Distinct Responses to Targeted Inhibitor Classes

Additionally, we examined *ex vivo* drug responses from the BEAT AML cohort used in our study, analyzing their associations with individual NBS clusters. We observed significant variability in response to multiple inhibitor families across NBS-defined clusters. A total of 34 drugs showed statistically significant differences in response across eight unique cluster pairs, highlighting the therapeutic heterogeneity among these molecular subtypes (Supplementary table1). The complete nomenclature of drug families is provided in Supplementary Table 2.

Despite the predominance of *FLT3* mutations in both clusters, Cluster 4 showed significantly greater sensitivity than Cluster 5 to targeted therapies inhibiting FLT3, VEGF, and the PI3K/AKT/mTOR pathways. In direct comparisons, Cluster 4 exhibited enhanced responses to quizartinib (RTK_TYPE_III, p = 0.021), sorafenib (RTK_TYPE_III/VEGFR/RET, p = 0.025), dovitinib (RTK_TYPE_III, p = 0.0002), axitinib (RTK_TYPE_III/VEGFR, p = 0.037), and A-674563 (PI3K/AKT/mTOR, p = 0.007). In contrast, Cluster 5 was more sensitive to the JAK inhibitor ruxolitinib (JAK, p = 0.042), indicating differential pathway dependencies between the two clusters (Figure 3A-D). Cluster 4 also exhibited more sensitivity than Cluster 3, showing the most extensive differences with significant responses to a broad range of inhibitors, including alisertib (AURK, p = 0.014), AZD1480 (JAK, p = 0.027), A-674563 (PI3K/AKT/mTOR, p = 0.036), and erlotinib (RTK_ERBB, p = 0.007). However, selumetinib (AZD6244), which targets MEK and the RAF/MEK/ERK pathway, was an exception, showing greater sensitivity in Cluster 3 (p = 0.043) (Supplementary Table 1). These findings may reflect that Cluster 4 is more broadly sensitive to targeted therapies than Clusters 3 and 5, potentially. L due to its high frequency of mutations—*FLT3* (100%), *NPM1* (100%), *DNMT3A* (65%), and *TET2* (39%). In contrast, Cluster 5, despite a predominance of *FLT3* mutations (96%), lacks *NPM1* mutations while *DNMT3A* mutations (20%) were less frequent, potentially limiting response of the Cluster 5 samples to the tested inhibitors. Cluster 3 shows a more distributed mutation profile without dominant targets, which may reduce its sensitivity to specific drug classes. The concentration of multiple targetable mutations in Cluster 4 likely underlies its enhanced therapeutic responsiveness.

**Figure 3.**
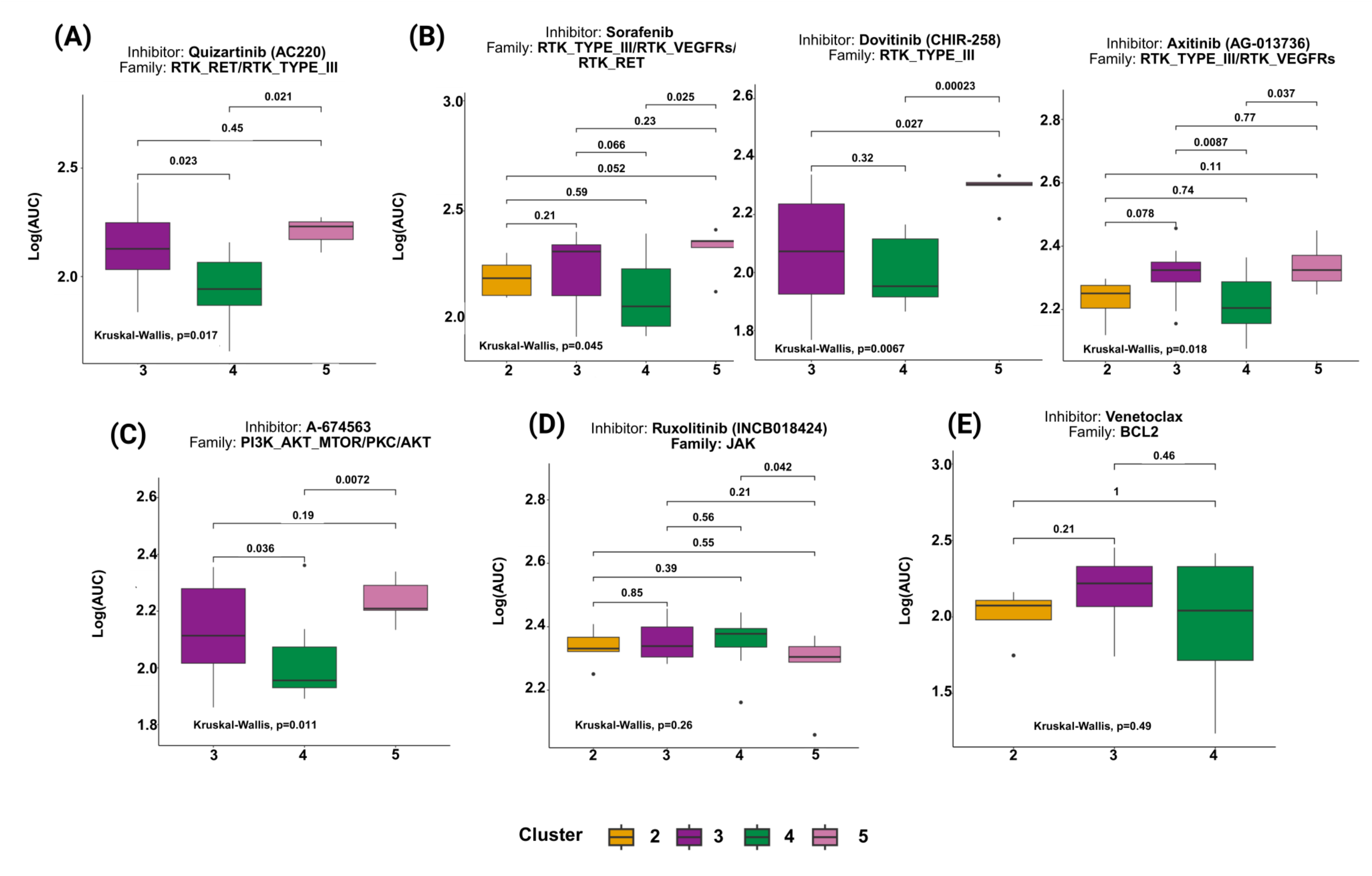
NBS-defined AML subtypes exhibit distinct ex vivo drug responses in the BEAT AML dataset. Lower values log (AUC) indicates better response to treatment. For drug family abbreviations, see Supplementary Table 2. **A)** Quizartinib (FLT3 inhibitor) showed better activity in Cluster 4 (*FLT3*-ITD, *NPM1* and *DNMT3A* mutation group) among the 5 clusters. **B)** Inhibitors sorafenib, dovitinib, axitinib that targets receptor tyrosine kinases (RTKs) III and RTK_VEGFRs showed better activity in Cluster 4. **C)** A-674563, which targets MTOR/AKT, was more active in Cluster 4 than Cluster 5. **D)** Ruxolitinib, a JAK inhibitor, showed significantly better activityin Cluster 5 than Cluster 4. **E)** Response to venetoclax was not significantly different among Clusters 2, 3 and 4.

Meanwhile, Cluster 2 showed differential sensitivity based on Kruskal-Wallis p-values, particularly to ERBB and ABL inhibitors, with significantly greater responses to lapatinib (RTK_ERBB, p = 0.02), erlotinib (RTK_ERBB, p = 0.07), and nilotinib (ABL, p = 0.025), suggesting alternative targeted strategies for this genetically distinct subgroup enriched in *DNMT3A* and *IDH2* mutations (Supplementary Figure 2A-B). The BCL2 inhibitor venetoclax showed no significant difference in response, suggesting comparable apoptotic pathway activity. These findings suggest that specific clusters harbor unique vulnerabilities to targeted therapies, particularly within the RTK, JAK, AURK, MEK, and PI3K/AKT/mTOR pathways. This supports the potential for personalized treatment strategies based on NBS-defined molecular subtypes in intermediate-risk AML.

### NBS-Defined AML Clusters Exhibit Distinct Transcriptomic and Pathway Signatures

To define the molecular features and pathway alterations distinguishing each cluster, we performed differential gene expression analysis using DESeq2^20^ followed by pathway enrichment analysis using fgsea^21^ and GO term enrichment using clusterProfiler^22^, based on the BEAT AML dataset^16^. We focused on identifying transcriptomic signatures and enriched biological processes that characterize each cluster in comparison to others. Log fold changes and adjusted p-values for differentially expressed genes from cluster-specific RNA-seq analyses are provided in Supplementary Table 3.

To understand the differential drug response between Cluster 4 and Cluster 5, we aimed to identify specific molecular alterations distinguishing these two subgroups. Differential expression analysis between Cluster 4 and Cluster 5 identified 74 significantly upregulated and 51 downregulated genes in Cluster 4 (log₂FC > 2 or < –2, adj. p < 0.05). Upregulated genes included *HLF*, *CXCL8* (IL-8), and *HES1*, while downregulated genes included *MECOM* (EVI1), *GAS6*, and *NDRG2* (Figure 4A). Pathway enrichment analysis revealed that Cluster 4 was positively enriched for inflammatory and immune-related pathways, including TNFα signaling via NF-κB (NES = 2.69), interferon gamma response (NES = 1.95), and TGF-β signaling (NES = 1.93), all with adjusted p-values < 0.001. This inflammatory signaling profile may underlie the heightened sensitivity of Cluster 4 to VEGFR, multiple RTK pathway and AKT pathway inhibitors^28,29^, with the notable exception of a non-specific JAK inhibitors (Figure 3). In contrast, proliferative and metabolic pathways such as MYC targets (NES = –3.02), E2F targets (NES = –3.48), and oxidative phosphorylation (NES = –2.96) were negatively enriched (Figure 4B). Additionally, biological process-level GSEA showed positive enrichment of positive regulation of leukocyte migration (NES = 1.99, adj. p = 0.0042) and response to glucagon NES = 1.90, adj. p = 0.0020), and negative enrichment of regulation of DNA-templated transcription (NES = -1.83, adj. p = 0.0023) and RNA biosynthetic processing (NES = -1.82, adj. p = 0.0023), highlighting distinct transcriptional programs between the two clusters, which could potentially be mediated by mutations in *NPM1* and *DNMT3A* (Figure 4C). However, when comparing Cluster 4 to all other patients, we observed significant negative enrichment of multiple pathways including those related to metabolism (e.g., glycolysis, adipogenesis, xenobiotic metabolism, oxidative phosphorylation), and immune response (e.g., interferon alpha/gamma response, inflammatory response). (Supplementary Figure 3A and 3B). Notably, several of these immune-related pathways were positively enriched in the Cluster 4 *vs*. Cluster 5 comparison, likely due to differences in the reference group affecting enrichment direction and aligning with the pronounced differences in drug sensitivity observed between the two clusters (Figure 3).

**Figure 4.**
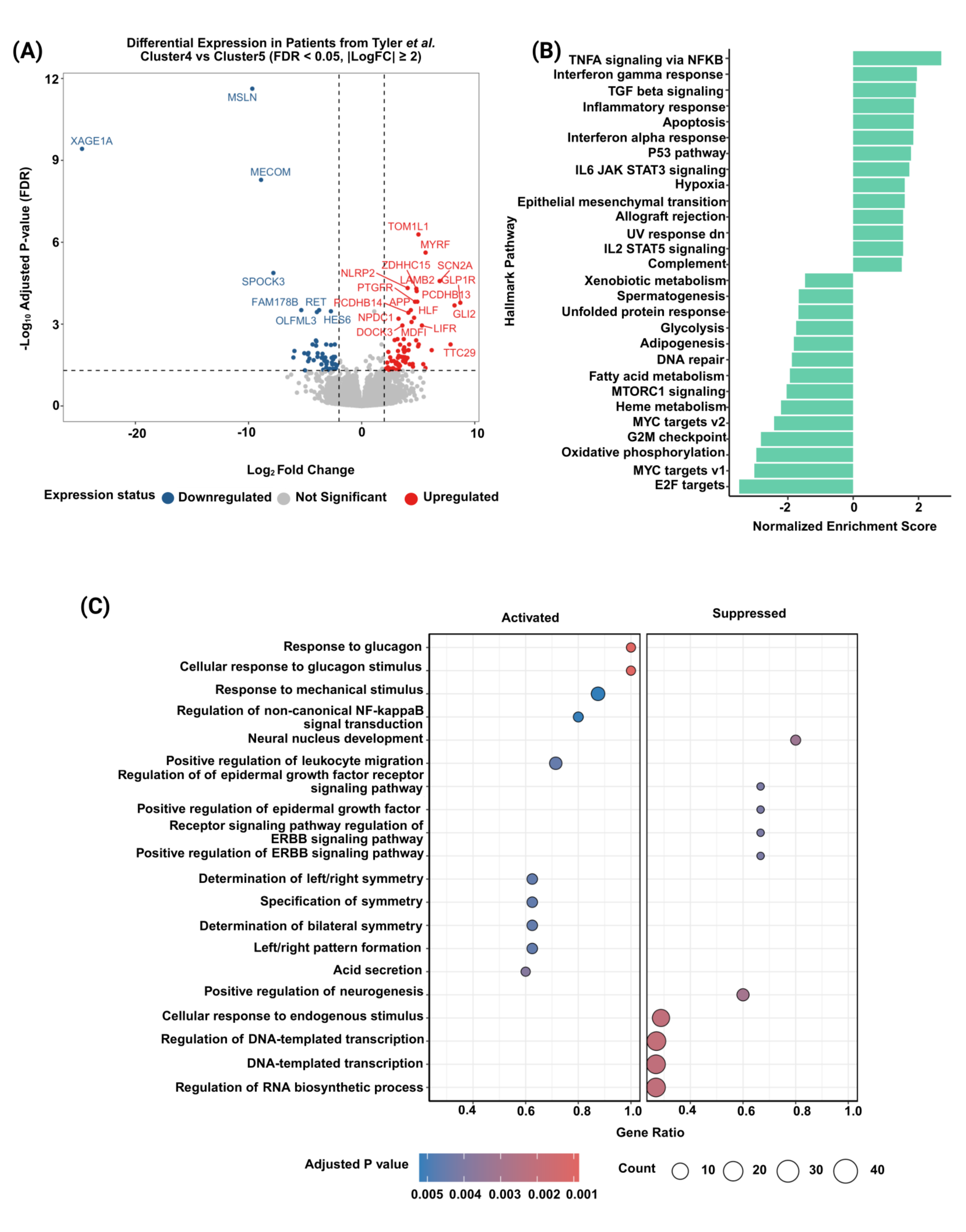
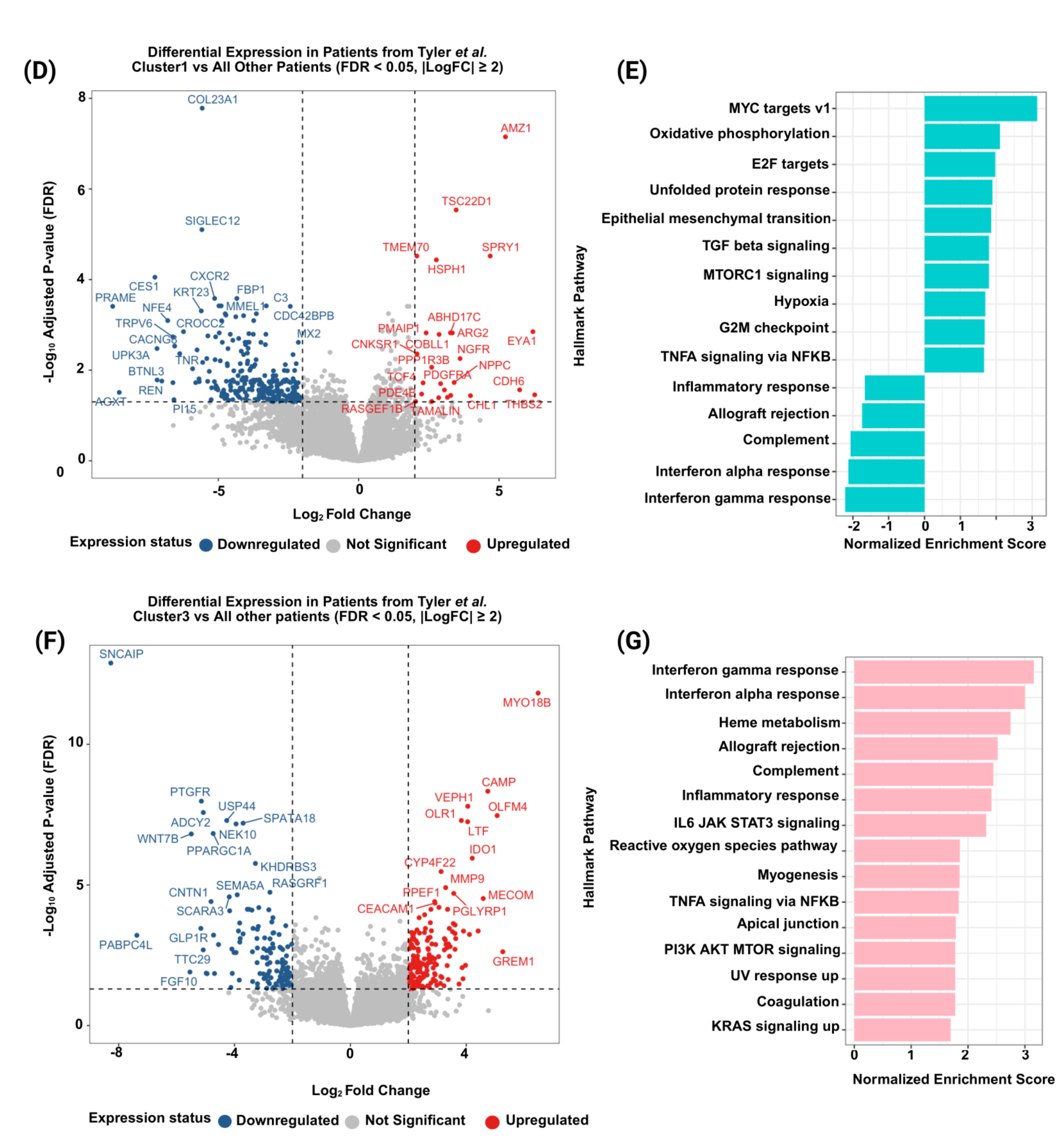
Distinct transcriptomic landscapes and pathway signatures characterize NBS-defined AML clusters. **A)** Differential gene expression analysis was performed on the BEAT AML dataset between patients assigned to Cluster 4 (*FLT3*-ITD, *NPM1* and *DNMT3A* mutations) versus Cluster 5 (*FLT3* mutation alone) as reference. An FDR cut-off of 0.05 and |LogFC| ≥ 2 was used to select differentially expressed genes. **B)** Pathway enrichment analysis was performed using the fast gene set enrichment analysis (fgsea) method. Ranked gene statistics were derived from differential expression analysis comparing Cluster 4 *vs*. Cluster 5 (reference) and hallmark pathways were used for enrichment. The pathways with an adjusted p-value (padj) < 0.01 were considered significantly enriched. Hallmarks pathways related to inflammation showed positive enrichment in Cluster 4 as compared to Cluster 5, whereas MYC target pathways are negatively enriched in Cluster 4. **C)** Biological significance of differentially expressed genes was assessed using Gene Set Enrichment Analysis (GSEA) for Gene Ontology (GO) biological processes. Go terms related to ERBB signaling pathway and epidermal growth factor were suppressed in Cluster 4. **D)** Differential gene expression analysis was performed on patients assigned to Cluster 1 (*IDH1* and D*NMT3A* mutations) versus All other patients (reference) in the BEAT AML dataset. E) Pathway enrichment analysis (fgsea method) using differentially expressed gene set in cluster 1 showed down-regulation of inflammatory pathway compared to all other patients. **F)** Differential gene expression analysis was performed on patients assigned to Cluster 3 (rare CH mutations) versus All other patients (reference) in the BEAT AML dataset used in our study. **G)** Pathway enrichment analysis (fgsea method) using differentially expressed genes in Cluster 3 revealed up-regulation of inflammatory pathways compared to all other patients.

Differential expression analysis of Cluster 1 against all other clusters revealed a distinct transcriptional profile with upregulation of genes such as *BAALC*, *PDGFRA*, and *PMAIP1*, and downregulation of *PRAME*, *MECOM*, *LCN2*, *HLF*, *SAMHD1*, *CXCR1/2/3*, *SIGLEC*s, and *G0S2*. Pathway enrichment analysis of Cluster 1 (adjusted p-value < 0.01) demonstrated a transcriptional program marked by upregulation of proliferative and metabolic pathways, including MYC targets (NES = 3.14), oxidative phosphorylation (NES = 2.11), and E2F targets (NES = 1.97), consistent with the functional role of *IDH1* mutations. In contrast, immune-related pathways such as complement activation (NES = –2.06), interferon alpha response (NES = –2.12), interferon gamma response (NES = –2.20), inflammatory response (NES = –1.66), and allograft rejection (NES = –1.74) were significantly downregulated, suggesting suppression of immune signaling (Figure 4D and E). Cluster 2, which was dominantly characterized by *DNMT3* mutations, exhibited minimal differential gene expression relative to other clusters—likely reflecting the widespread presence of *DNMT3* mutations across the cohort, which may have reduced transcriptional contrasts between groups (Supplementary Figure 3E).

In Cluster 3, genes such as *MECOM*, *CEACAM1*, *CEACAM5*, *CEACAM6*, *MMP9*, *IL6*, *IL27*, *CD274*, *ITGB3*, and *LCN2* were upregulated compared to all other clusters. Conversely, *PPARGC1A*, *KITLG*, *UCHL1*, *WNT7B*, and *WNT9A* were downregulated. Pathway enrichment analysis indicated a strong immune-activated transcriptional profile, with significant positive enrichment of interferon gamma response (NES = 3.16), interferon alpha response (NES = 3.00), and inflammatory response (NES = 2.41). These findings are consistent with previously reported enrichment of inflammatory pathways mediated by clonal hematopoiesis^30^. Additional enrichment in heme metabolism (NES = 2.76) and IL6-JAK-STAT3 signaling (NES = 2.32) may reflect altered erythroid or myeloid differentiation (Figure 4F and G). Compared to all other clusters, Cluster 5 displayed a distinct transcriptional profile with upregulation of *RET*, *RSPO1*, *FGL1*, and *LXN,* and downregulation of *OLFM4*, *LCN2*, *CDH2*, *HLF*, *THBS1*, *ESAM*, *CXCL8*, *CXCL3*, and *DPP4*. fgsea analysis (adjusted p-value < 0.01) showed enrichment of proliferative and metabolic pathways, including E2F targets (NES = 3.65), MYC targets V1 (NES = 2.96), and V2 (NES = 2.93). In contrast, immune and inflammatory pathways such as TNFα signaling via NF-κB (NES = -2.80), inflammatory response (NES = -2.45), and interferon gamma response (NES = 2.45) were significantly downregulated (Supplementary Figure 3C and 3D). These findings suggest that Cluster 5 is highly proliferative and metabolically active, with suppressed immune signaling, likely mediated by the predominant *FLT3* mutations.

## DISCUSSION

This analysis presents a ML approach that deconvolutes the heterogeneity within intermediate-risk AML patients, revealing subgroups with distinct clinical outcomes and molecular characteristics. Leveraging the NBS method, which integrates molecular pathway networks with mutation data, this unsupervised approach detects common pathways affected by diverse mutations through network propagation. We utilized a cancer-specific network composed exclusively of high-confidence interactions related to cancer, incorporating genetic alterations associated with AML. This enabled a more robust classification of tumors into clinically relevant subtypes. Ultimately, this framework delineated distinct patient subsets with unique mutational landscapes, functional profiles, and survival outcomes. Notably, we uncovered significant heterogeneity within the intermediate-risk AML group defined by the ELN 2022 classification. Using NBS, we identified a subgroup with inferior clinical outcomes and distinct molecular features characterized by transcriptional dysregulation.

Cluster 4, in particular, exhibited the poorest survival among all intermediate-risk clusters (Figure 2), whereas other clusters showed comparable outcomes. This high-risk subgroup was enriched for *NPM1* and *FLT3*-ITD mutations, along with clonal hematopoiesis-associated mutations in *DNMT3A* and *TET2*. They also demonstrated elevated blast counts and lacked additional chromosomal abnormalities. Notably, these adverse clinical outcomes were not explained by differences in initial response to chemotherapy or stem cell transplant (Supplementary Figure 1). The molecular features of Cluster 4 align with previous findings in *NPM1*-mutated AML, where co-mutations in *FLT3*-ITD and *DNMT3A* increase measurable residual disease (MRD) positivity and relapse risk, even after MRD negativity ^31^. These findings reinforce the distinct biological identity of Cluster 4 as defined by the NBS-ML framework.

Cluster 4 also differed significantly from Cluster 5 AMLs, which had *FLT3* mutations but lacked concurrent *NPM1* and *DNMT3A*/*TET2* mutations. For instance, in *ex vivo* drug sensitivity assays (Figure 3), Cluster 4 responded more favorably to quizartinib than Cluster 5, suggesting that the combination of *FLT3*-ITD with *NPM1* and *DNMT3A* mutations may enhance sensitivity to FLT3 inhibitors. This observation mirrors results from the QuANTUM-First phase 3 trial ^32^. Additionally, Cluster 4 showed improved responses to VEGFR and AKT inhibitors compared to Clusters 3 and 5, while Cluster 5 was more sensitive to the JAK inhibitor ruxolitinib. Venetoclax responses were similar across Clusters 2, 3, and 4 (Figure 3), suggesting that *NPM1* co-mutations do not necessarily confer enhanced sensitivity to venetoclax, consistent with prior clinical observations ^33^. These data suggest that the molecular and protein network alterations in Cluster 4 may lead to specific pathway activations—particularly in RTK/VEGFR and AKT signaling—that are amenable to targeted therapy. Therefore, a more selective biomarker-driven strategy focused on patients with *NPM1*, *FLT3*-ITD, and CH mutations (e.g., Cluster 4) may offer greater therapeutic benefit than trials using unselected patient cohorts ^34,35^. Transcriptomic profiling further supported the *ex vivo* drug sensitivity data (Figures 3 and 4). These findings illustrate how ML-based stratification, integrating mutational and pathway-level data, can uncover biologically and clinically meaningful patient subgroups.

Distinct dominant mutations were observed in most clusters, except Cluster 3, even though the total mutation burden was similar across groups. For example, *IDH1* mutations characterized Cluster 1, *DNMT3A* defined Cluster 2, and *FLT3* characterized Cluster 5. These patterns point to clonal evolution driven by specific mutations, resulting in unique pathway activations. Cluster 1, enriched for *IDH1* mutations, showed mitochondrial pathway activation and reduced inflammatory signaling, consistent with the biological function of *IDH1* mutations. In contrast, Cluster 3 comprised AML cases with diverse mutations typical of clonal hematopoiesis or MDS-like features, often linked with therapy-related or transformed AMLs (Figure 2). This cluster was characterized by enrichment of inflammatory signaling pathways, which are progressively activated during the transition from clonal hematopoiesis to overt myeloid malignancy ^36^ (Figure 4). Interestingly, Cluster 2, defined by *DNMT3A* mutations, lacked distinct pathway enrichment. This may be due to *DNMT3A*’s broad role as a founding mutation in AML, contributing to a shared molecular baseline across clusters (Supplementary Figure 3).

Despite the valuable insights generated by this ML–based framework, several limitations must be acknowledged. First, this analysis was conducted on a relatively small patient cohort, which may limit the statistical power and generalizability of the findings. Larger, more diverse cohorts will be necessary to validate the robustness of the identified clusters and their associated clinical outcomes. Second, while the NBS method effectively integrates mutation data with pathway-level interactions, its performance is inherently dependent on the quality and comprehensiveness of the underlying interaction network, which may miss rare or context-specific interactions. Additionally, this framework focuses primarily on somatic mutations and protein-protein interaction networks, without incorporating additional layers of molecular complexity, such as epigenetic alterations, non-coding RNA profiles, or immune landscape, which could further refine patient stratification. Finally, the therapeutic predictions derived from pathway activation and literature-based associations require prospective clinical validation to confirm their clinical utility and translational potential.

In summary, our findings reveal substantial heterogeneity within intermediate-risk AML under the ELN 2022 criteria. The NBS-based ML framework effectively stratifies AML cases by integrating mutational and protein network data, identifying subgroups with differing prognoses and treatment sensitivities. Our approach, developed using unsupervised ML techniques, is compatible with standard clinical mutation panels and offers a promising strategy for enhancing risk assessment and tailoring therapy. The identification of a high-risk subgroup with actionable molecular features highlights opportunities for targeted interventions and supports the use of genetic profiles to define treatment-eligible populations. This ML-driven framework may pave the way for hypothesis generation and personalized therapeutic development targeting specific resistance mechanisms.

## Data Availability

All data produced in the present study are available upon reasonable request to the authors

## ACKNOWLEDGEMENTS

The authors are grateful for the generous donation of samples from the contributing patients. The samples and clinical data for the FIMM cohort were obtained from the Finnish Hematology Registry and Biobank. The authors also thank the FIMM Genomics Unit, which is supported by HiLIFE and Biocenter Finland. This work has been supported by funding the Research Council of Finland (grant no. 334781, 352265, 357686, and 320185), the Sigrid Jusélius Foundation, the Cancer Foundation Finland and the American Cancer Society to support this study. We acknowledge the use of ChatGPT for text formatting and language polishing. The authors also acknowledge the use of BioRender.com for the generation of figure illustrations.

## AUTHORSHIP CONTRIBUTIONS

JW, MVK and CAH contributed to the conception/design of the study. Data analysis was performed by AS, JS, PS with advice from all authors. JW & AS wrote the manuscript. All authors critically reviewed and contributed to the final manuscript.

