## Supplementary Figures for "Network-Based Stratification Refines Stratification of Intermediate-Risk Acute Myeloid Leukemia Samples"

### Slide 1
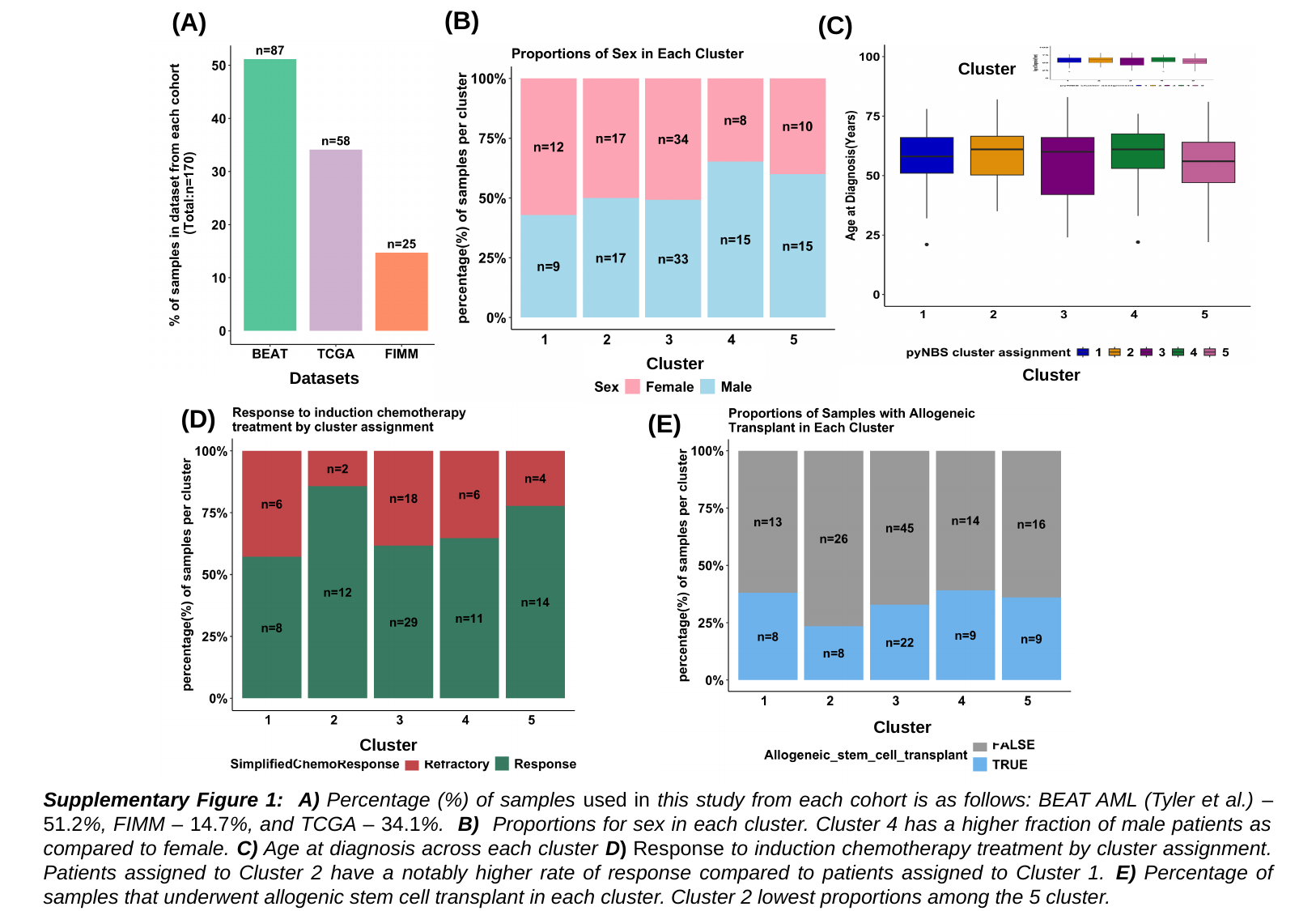

(B)
(A)
Cluster
Cluster
Cluster
(D)
Cluster
(E)
(C)
Datasets
Cluster
Supplementary Figure 1: A) Percentage (%) of samples used in this study from each cohort is as follows: BEAT AML (Tyler et al.) – 51.2%, FIMM – 14.7%, and TCGA – 34.1%. B) Proportions for sex in each cluster. Cluster 4 has a higher fraction of male patients as compared to female. C) Age at diagnosis across each cluster D) Response to induction chemotherapy treatment by cluster assignment. Patients assigned to Cluster 2 have a notably higher rate of response compared to patients assigned to Cluster 1. E) Percentage of samples that underwent allogenic stem cell transplant in each cluster. Cluster 2 lowest proportions among the 5 cluster.

### Slide 2
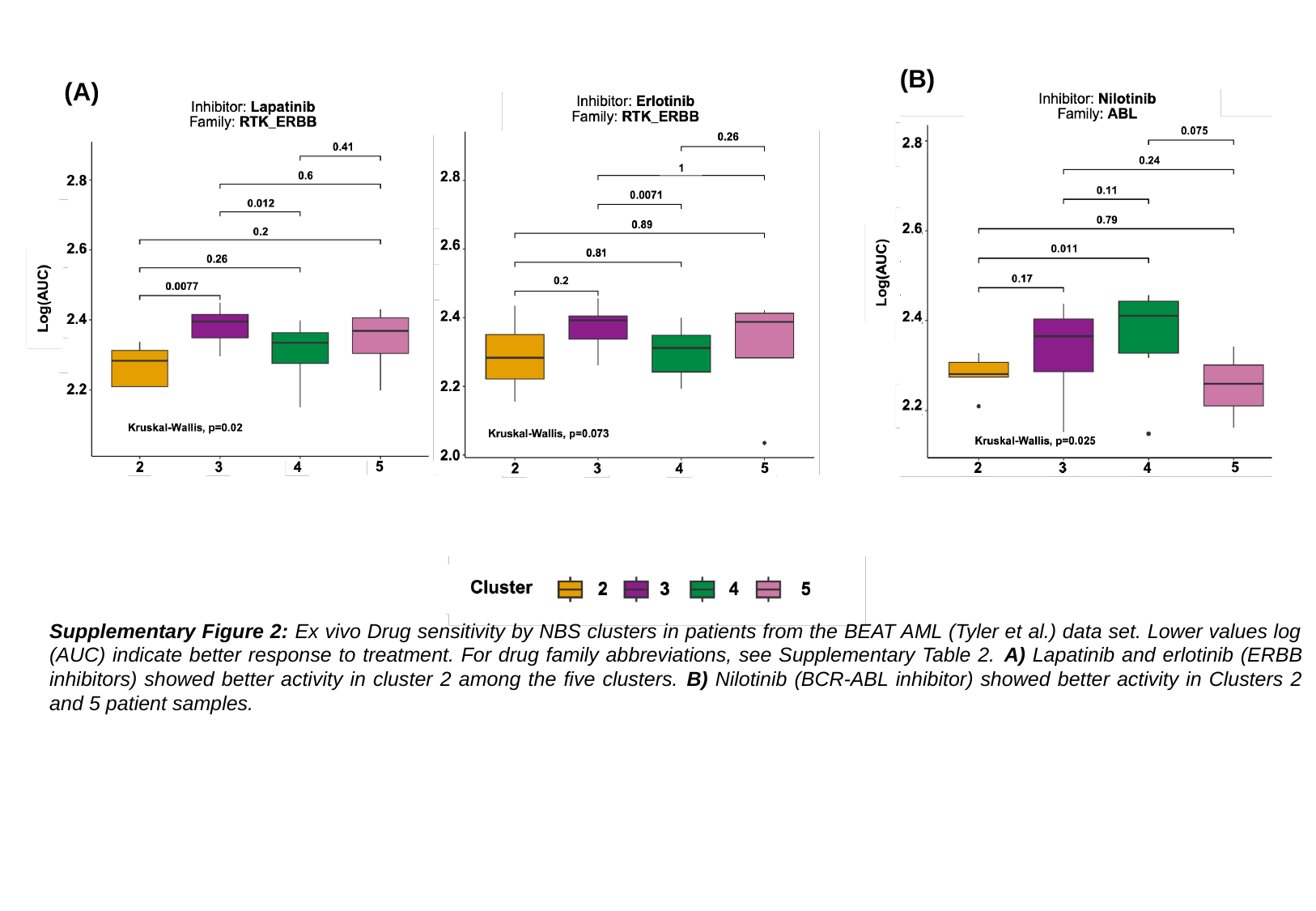

(B)
(A)
Supplementary Figure 2: Ex vivo Drug sensitivity by NBS clusters in patients from the BEAT AML (Tyler et al.) data set. Lower values log (AUC) indicate better response to treatment. For drug family abbreviations, see Supplementary Table 2. A) Lapatinib and erlotinib (ERBB inhibitors) showed better activity in cluster 2 among the five clusters. B) Nilotinib (BCR-ABL inhibitor) showed better activity in Clusters 2 and 5 patient samples.

### Slide 3
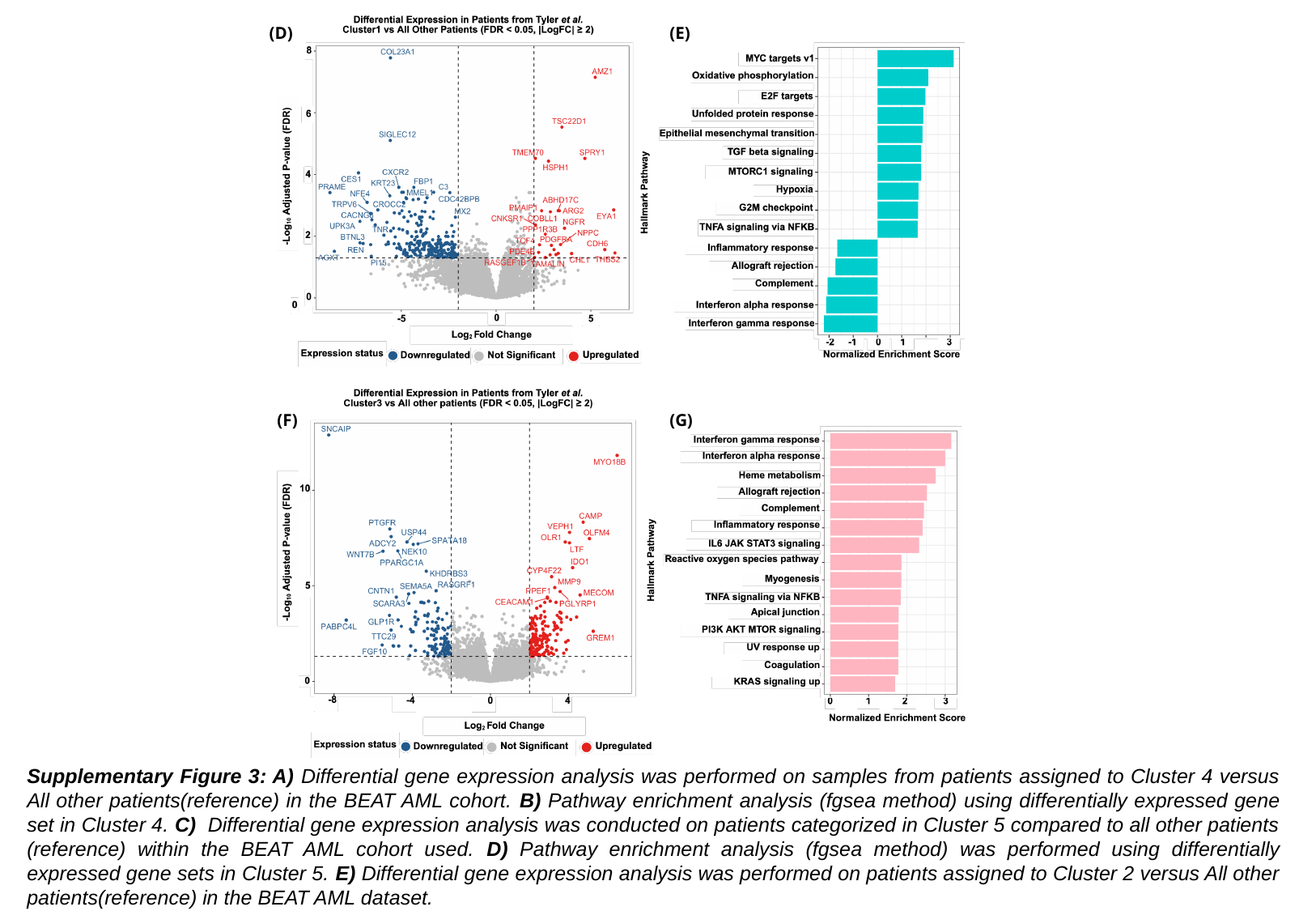

(E)
(D)
(G)
(F)
Supplementary Figure 3: A) Differential gene expression analysis was performed on samples from patients assigned to Cluster 4 versus All other patients(reference) in the BEAT AML cohort. B) Pathway enrichment analysis (fgsea method) using differentially expressed gene set in Cluster 4. C) Differential gene expression analysis was conducted on patients categorized in Cluster 5 compared to all other patients (reference) within the BEAT AML cohort used. D) Pathway enrichment analysis (fgsea method) was performed using differentially expressed gene sets in Cluster 5. E) Differential gene expression analysis was performed on patients assigned to Cluster 2 versus All other patients(reference) in the BEAT AML dataset.
