## Supplementary Table 1-2 for "Network-Based Stratification Refines Stratification of Intermediate-Risk Acute Myeloid Leukemia Samples"

**Supplementary Table1:** Pairwise T-Tests Between Clusters for Each Inhibitor Group: Eight unique cluster pairs with significant p-values for 34 drugs

| **Inhibitor** | **Cluster pair** | **P Value** | **Drug target Family** |
| --- | --- | --- | --- |
| Crenolanib | 3,2 | 0,038 | RTK_TYPE_III |
| Lapatinib | 3,2 | 0,008 | RTK_ERBB |
| Lenalidomide | 3,4 | 0,035 | n/a |
| Tozasertib (VX-680) | 3,4 | 0,046 | AURK; RTK_TYPE_III; RTK_RET |
| Nilotinib | 4,2 | 0,011 | ABL |
| Ponatinib (AP24534) | 4,2 | 0,029 | RTK_FGFRs; RTK_VEGFRs; RTK_EPH; RTK_TYPE_III; RTK_RET; SFKs; RTK_ALK_MET; ABL |
| A-674563 | 4,3 | 0,036 | PI3K_AKT_MTOR; PKC; AKT |
| Alisertib (MLN8237) | 4,3 | 0,014 | AURK |
| Axitinib (AG-013736) | 4,3 | 0,009 | RTK_TYPE_III; RTK_VEGFRs |
| AZD1480 | 4,3 | 0,027 | JAK; BTK_TEC; RTK_NTRK |
| Bosutinib (SKI-606) | 4,3 | 0,018 | BTK_TEC; ABL; SFKs |
| Canertinib (CI-1033) | 4,3 | 0,025 | RTK_ERBB |
| CHIR-99021 | 4,3 | 0,016 | GSK3 |
| Crizotinib (PF-2341066) | 4,3 | 0,051 | RTK_ALK_MET; RTK_TAM; RTK_NTRK |
| Erlotinib | 4,3 | 0,007 | RTK_ERBB |
| Gefitinib | 4,3 | 0,052 | RTK_ERBB |
| GSK-1838705A | 4,3 | 0,027 | RTK_INSR_IGF1R; RTK_ALK_MET |
| GSK690693 | 4,3 | 0,020 | PKC; PI3K_AKT_MTOR; AKT |
| KW-2449 | 4,3 | 0,006 | BTK_TEC; RTK_TYPE_III; AURK |
| Lapatinib | 4,3 | 0,012 | RTK_ERBB |
| MLN120B | 4,3 | 0,029 | IkK |
| NVP-ADW742 | 4,3 | 0,013 | RTK_INSR_IGF1R |
| NVP-TAE684 | 4,3 | 0,015 | RTK_TAM; FAKs; RTK_ALK_MET; BTK_TEC; RTK_INSR_IGF1R; RTK_TIE |
| Quizartinib (AC220) | 4,3 | 0,023 | RTK_RET; RTK_TYPE_III |
| Roscovitine (CYC-202) | 4,3 | 0,019 | pan_CDK |
| Selumetinib (AZD6244) | 4,3 | 0,043 | MEK; RAF_MEK_ERK |
| SGX-523 | 4,3 | 0,002 | RTK_ALK_MET |
| Vargetef | 4,3 | 0,013 | n/a |
| A-674563 | 4,5 | 0,007 | PI3K_AKT_MTOR; PKC; AKT |
| Afatinib (BIBW-2992) | 4,5 | 0,020 | RTK_ERBB |
| Axitinib (AG-013736) | 4,5 | 0,037 | RTK_TYPE_III; RTK_VEGFRs |
| JNJ-7706621 | 4,5 | 0,053 | pan_CDK; AURK; RTK_TIE |
| KI20227 | 4,5 | 0,034 | RTK_TYPE_III |
| KW-2449 | 4,5 | 0,017 | BTK_TEC; RTK_TYPE_III; AURK |
| PHT-427 | 4,5 | 0,044 | n/a |
| Quizartinib (AC220) | 4,5 | 0,021 | RTK_RET; RTK_TYPE_III |
| Ruxolitinib (INCB018424) | 4,5 | 0,042 | JAK |
| Sorafenib | 4,5 | 0,025 | RTK_TYPE_III; RTK_VEGFRs; RTK_RET |
| Sorafenib | 5,2 | 0,052 | RTK_TYPE_III; RTK_VEGFRs; RTK_RET |
| Dovitinib (CHIR-258) | 5,3 | 0,027 | RTK_TYPE_III |
| Elesclomol | 5,3 | 0,038 | n/a |
| GSK-1838705A | 5,3 | 0,052 | RTK_INSR_IGF1R; RTK_ALK_MET |
| JAK Inhibitor I | 5,3 | 0,048 | JAK |
| TG100-115 | 5,3 | 0,005 | PI3K_MTOR; PI3K_AKT_MTOR |
| Dovitinib (CHIR-258) | 5,4 | 0,000 | RTK_TYPE_III |
| JAK Inhibitor I | 5,4 | 0,026 | JAK |

**Supplementary Table 2:** Abbreviations and Full Names of Drug Families present in the BEAT AML Dataset

| **Drug target Family** | **Full Name** |
| --- | --- |
| PKC | Protein Kinase C |
| AKT | AKT |
| PI3KAKTMTOR | PI3K/AKT/mTOR Pathway |
| CDK4_6 | Cyclin-Dependent Kinase 4/6 |
| pan_CDK | Pan Cyclin-Dependent Kinases |
| JAK | Janus Kinase |
| RTK_NTRK | Receptor Tyrosine Kinase NTRK |
| BTK_TEC | Bruton's Tyrosine Kinase/TEC |
| RTK_ERBB | Receptor Tyrosine Kinase ERBB |
| AURK | Aurora Kinase |
| RTK_VEGFRs | Receptor Tyrosine Kinase VEGFRs |
| RTKTYPEIII | Receptor Tyrosine Kinase Type III |
| PI3K_MTOR | PI3K/mTOR Pathway |
| PLK | Polo-Like Kinase |
| IkK | IκB Kinase |
| proteasome | Proteasome |
| SFKs | Src Family Kinases |
| ABL | Abelson Tyrosine Kinase |
| GSK3 | Glycogen Synthase Kinase 3 |
| MEK | Mitogen-Activated Protein Kinase |
| RAFMEKERK | RAF/MEK/ERK Pathway |
| RTK_TIE | Receptor Tyrosine Kinase TIE |
| RTK_RET | Receptor Tyrosine Kinase RET |
| RTKALKMET | Receptor Tyrosine Kinase ALK/MET |
| RTK_TAM | Receptor Tyrosine Kinase TAM |
| RTK_EPH | Receptor Tyrosine Kinase EPH |
| p38 | p38 Mitogen-Activated Protein Kinase |
| SYK | Spleen Tyrosine Kinase |
| RAF | RAF Kinase |
| RTKINSRIGF1R | Receptor Tyrosine Kinase INSR/IGF1R |
| BRD4 | Bromodomain-Containing Protein 4 |
| ATM | Ataxia Telangiectasia Mutated |
| FAKs | Focal Adhesion Kinases |
| mdm2 | Mouse Double Minute 2 |
| HDAC | Histone Deacetylase |
| RTK_FGFRs | Receptor Tyrosine Kinase FGFRs |
| CAMK | Calcium/Calmodulin-Dependent Kinase |
| BCL2 | B-Cell Lymphoma 2 |
| SHH | Sonic Hedgehog |
